# Estimate of the Maximum Limit of Total Cases of Infected Patients COVID-19

**DOI:** 10.1101/2020.04.10.20060822

**Authors:** Carlos Maximiliano Dutra, Carlos Augusto Riella de Melo

## Abstract

In this work, we present a method to estimate the maximum limit of total cases COVID-19 cases considering that the time in which the maximum number of new daily cases occurs corresponds to the inflection point of the curve described by the total number of cases that assumed to have a growth according to a logistical function in which the number of total cases at the inflection point will correspond to half of the maximum limit of total cases COVID-19. We estimate this maximum limit for China and South Korea, obtaining results compatible with the observations. And we also estimate for Italy, Germany, United Kingdom, United States and Spain.

## Introduction

According to World Health Organization - WHO (SR1, 2020), on December 31, 2019, the Chinese office at WHO reported cases of pneumonia of unknown cause in the city of Wuhan in Hubei Province in China; on January 20, 2020, there were already 258 cases in Hubei, being, on January 7, 2020, identified a new type of coronavirus (2019-nCoV). Since early February, WHO has officially called the disease caused by the new COVID-19 (Coronavirus Dease 2019), being the causative virus called SARS-CoV-2 (severe acute respiratory syndrome coronavirus 2) (Chan et al., 2020). On March 11, WHO (SR51, 2020) characterizes COVID-19 as a Pandemic. According to WHO report 78 (SR78, 2020), on April 7, 2020 the world has 1,279,722 cases, and 72,614 deaths from the disease.

Some work has appeared to estimate the number of infected cases, especially in Brazil, considering exponential growth models for the projection of cases of COVID-19 infection, among other authors we have: Figueiredo et al. (2020), Codeço et al. (2020) and Batista et. al (2020).

Prediction models are critical to simulate possible scenarios, and from these assess the need for significant changes in the flow of care for cases, increase the number of hospital beds, among other actions pertinent to the control and mitigation of the health problem considered. However, exponential growth models do not include the intermediate stage until the end of the epidemic, which goes from the tipping point with deceleration of exponential growth to the stability phase.

In the present study we present a methodology to determine a maximum limit for the total number of cases of patients infected with COVID-19.

## Theoretical foundation

The present estimate considers that the Total Number of COVID-19 infected patients registered with the World Health Organization (WHO – https://www.who.int/emergencies/diseases/novelcoronavirus-2019/situation-reports) over time presents a logistical growth with the phases as shown in Figure 1.

**Figure 1:**
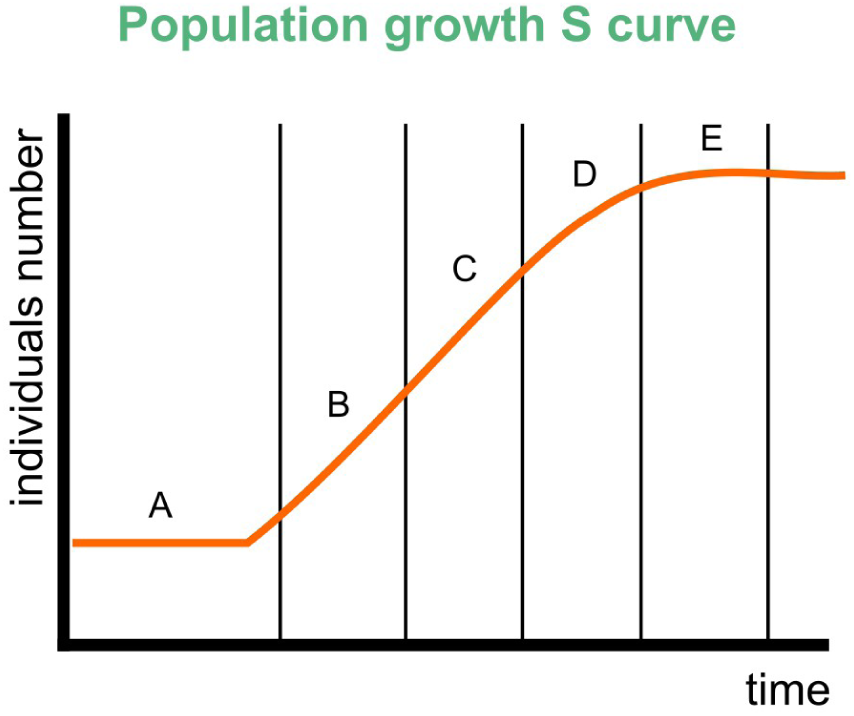
Description of the Logistic Growth Model The Logistic Model is characterized by a Sigmoidal growth-type function, an “S” shape. The exponential growth of the population suffers resistance from the environment. Stage A: slow growth, phase of adaptation of the population to the environment. Stage B: accelerated or exponential growth, Stage C: growth changes behavior becoming decelerated, the population is subject to the limits imposed by the environment, environmental resistance is higher on the population. Stage D: environmental resistance prevailing over exponential growth more intensely. Stage E: Asymptotic growth, in which the environmental resistance prevents from exceeding a certain maximum value. Source: http://matemabio.blogspot.com/p/dinamica-populacional.html

Bringing to our study variable, the Total Number of Cases presents: (a) slow growth at the beginning; (b) a phase of accelerated growth of the exponential type; (c) a phase of moderate exponential growth that is increasingly distancing itself from this model due to the slow growth; (d) an intermediate phase with greater growth deceleration and (e) a phase of stability – Plateau, where new cases gradually cease, with very slow and asymptotic growth.

Currently only two countries have reached stage E, China and South Korea, as shown in Figure 2. The variation in the total number of cases in the plateau region is attributed by China for the repatriation and coming of infected foreigners coming from abroad, the values of new cases are of a similar order between China and South Korea, however, the number of infected people in China is about 8 times greater so the difference in the plateau region for both countries. Nonetheless, we found that the logistic function can model the behavior of population growth in the total number of infected cases.

**Figure 2:**
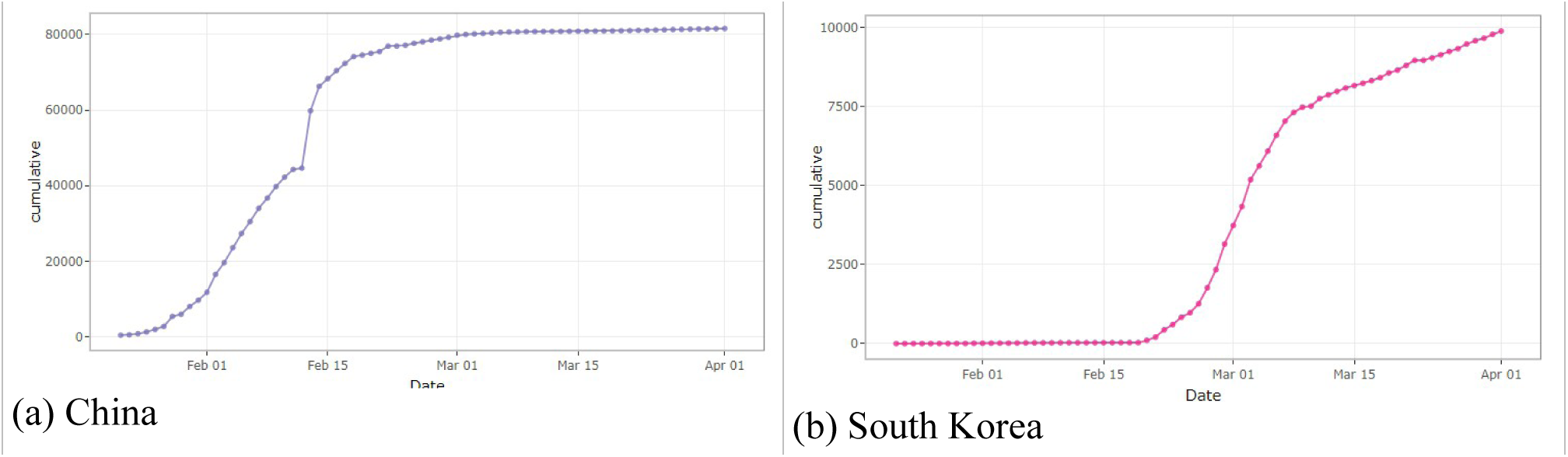
Total number of cases according to time Source: https://vac-lshtm.shinyapps.io/ncov_tracker/

Mathematically the logistic growth function is described by the equation below:

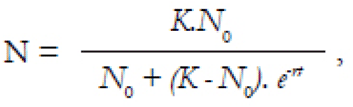

In which *N* is the number of individuals at time ^*t*^, *K* represents the population limit value, *N*_*0*_ is the initial population, ^*r*^ is the growth rate, and ^*t*^ is the time.

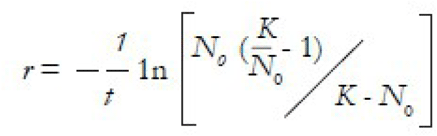

We can use the characteristics of the logistic function to estimate the Maximum Limit of Total Cases of COVID-19 infected patients, which will be defined by K the population limit value for the asymptotic behavior of the growth phase (e). This K value corresponds to Figure (3a) can be determined from the inflection point between the phases (b), of accelerated growth, and (c) of slowed growth of the growth curve, the y-coordinate value of the inflection point corresponds to K/2. Mathematically, the derivative or rate of change of the Logistic Function over time results in a function with a maximum peak that is associated with the inflection point (Figure 3b), and that function can be expressed by a Gaussian function to determine the time t at which the maximum occurs.

**Figure 3:**
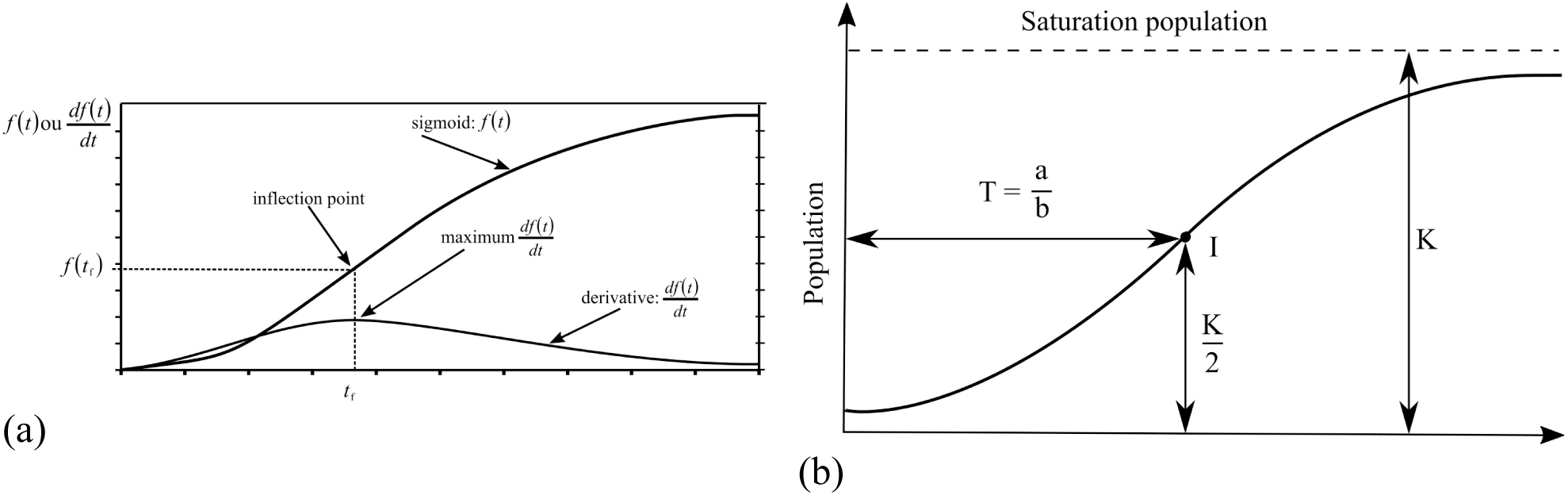
Graphical diagram of the Logistics function, the inflection point and its derivative.

The derivative of the total number of cases over time is nothing more than the number of new cases over time. Therefore, the tipping point of the function total number of cases occurs at the same time as the maximum number of new cases; and when determining the time t of the maximum number of new cases, we have the x coordinate to determine the total number of cases in the inflection that corresponds to the maximum limit of the total number of cases divided by 2.

## Methodology

Thus, we must access the World Health Organization website (https://www.worldometers.info/coronavirus/) and choose the country to be analyzed. We can read the graph of total number of cases by defining the initial date D0 and the total initial number of cases N0 as a reference for the model. Subsequently, we verify in the New Cases graph the date that the maximum number of new cases occurs DCNmax (if the maximum is not clear, it is necessary to perform a Gaussian function adjustment to determine the time t that occurs). Then we can see in the graph the total number of cases on this date DCNmax which is the value of the total number of cases NTC_DCmax, this is the value corresponding to the inflection point of this graph and thus the maximum limit value “K” corresponds to twice the NTC_DCNmax, allowing to know the limit of the total number of cases in that country. In order to be able to estimate by the Logistic Model, when the stability phase (Plateau) of the total number of cases, where there are practically very few new cases, will be reached we calculated the time interval Ti between the date of maximum new cases DCNmax and the initial reference date D0. From that time interval Ti, of the value of the maximum limit of total number of cases of COVID19 “K” patients and of the total number of initial cases of reference N0 it is possible to calculate the date by which, using the model, the total number of cases will be 90% K (D90%), 95% K(D95%), and 99% K (D99%).

In Table 1 we have the application of the method for China and South Korea that have already reached the growth stability phase (5), as well as for countries in Europe where the number of new cases has just reached.

**Table 1:** Synthesis of results using the methodology with updated data until 04/07/2020.

| Country | D0 | N0 | DCNmax | Ti | NTC_DCmax | K | D90% | D95% | D99% |
| --- | --- | --- | --- | --- | --- | --- | --- | --- | --- |
| China | 01/22/20 | 571 | 02/10/20 | 19<sup>a</sup> | 42638 | 85276 | 02/29/20 | 03/10/20 | 03/22/20 |
| SouthKorea | 02/15/20 | 28 | 03/03/20 | 17 | 5186 | 10372 | 03/17/20 | 03/25/20 | 04/08/20 |
| Italy | 01/29/20 | 3 | 03/26/20 | 61<sup>a</sup> | 80589 | 161178 | 04/03/20 | 04/15/20 | 05/08/20 |
| German | 02/15/20 | 16 | 03/30/20 | 44 | 66885 | 133770 | 04/18/20 | 05/05/20 | 06/05/20 |
| UK | 02/15/20 | 9 | 04/05/20 | 50 | 47806 | 95612 | 04/26/20 | 05/15/20 | 06/19/20 |
| USA | 02/15/20 | 15 | 04/04/20 | 49 | 311357 | 622714 | 04/19/20 | 05/07/20 | 06/07/20 |
| Spain | 02/15/20 | 2 | 04/01/20 | 46<sup>a</sup> | 87956 | 175912 | 04/19/20 | 05/06/20 | 06/06/20 |

<sup>a</sup> determined by adjusting a Gaussian function the data distribution of the number of new cases as a function of time.

For the cases of China and South Korea it is already possible to compare the values predicted by the model with the actual values. On D99% date = 03/22/20 the total number of cases recorded in China was 81,093 cases, whereas by the model presented we would have 99% of 85,276n = 84,423 cases, thus being possible to estimate a relative error of approximately 4%. As of date D95% = 03/25/20 the total number of cases recorded in South Korea was 9,137 cases, whereas by the model presented we would have 95% of 10,372 = 9,853 cases, giving a relative error of approximately 8%.

## Final considerations

Through this study it was possible to estimate the maximum number of cases of patients infected with COVID-19 from the definition of the maximum number of new cases, estimating when the plateau stability phase will occur one to two months in advance of this phase. This is an important variable for the development of prediction models that aim to estimate the impact of COVID-19 on the Brazilian Health System. It is intended to use the methodology to define this parameter in data from Brazil/States/Cities and also in other countries in the world during the Pandemic.

## Data Availability

The necessary data and sources are all contained in the manuscript.

## Notes

### Competing Interest Statement

The authors have declared no competing interest.

### Funding Statement

No external funding.

## References

Batista, et al. https://sites.google.com/prod/view/nois-pucrio/publica%C3%A7%C3%B5es#h.niiouup8rvuu

Chan, J.F.W. et al. A familial cluster of pneumonia associates with the 2019 novel coronavirus indicating person-to-person transmission: a study of a family cluster. The Lancet, v. 395, m. 10223, p. 514–523. 2020.

Codeço, C.T. et al. Estimativa de risco de espalhamento da COVID-19 nos estados brasileiros e avaliação da vulnerabilidade socioeconômica nos municípios. Relatório no 3 do Grupo de Métodos Analíticos de Vigilância Epidemiológica (MAVE), PROCC/Fiocruz e EMap/FGV, 2 de abril de 2020, http://covid-19.procc.fiocruz.br/

https://www.arca.fiocruz.br/bitstream/icict/40509/5/Relatorios_tecnico_COVID-19_procc-emap-covid-19-reporte3_resultados.pdf [acessado em 07 de abril de 2020]

Figueiredo, D. et. al COVID19 em dados: Brasil em perspectiva comparada. 2020. https://www.researchgate.net/profile/Dalson_Figueiredo2/publication/340278851_COVID-19_EM_DADOS_BRASIL_EM_PERSPECTIVA_COMPARADA/links/5e81e6eb92851caef4acfc6f/COVID-19-EM-DADOS-BRASIL-EM-PERSPECTIVA-COMPARADA.pdf [acessado em 07 de abril de 2020]

World Health Organization. Novel Coronavirus (2019-nCoV) Situation Report – 1. Janeiro 2020. https://www.who.int/docs/default-source/coronaviruse/situation-reports/20200121-sitrep-1-2019-ncov.pdf?sfvrsn=20a99c10_4 [acessado em 07 de abril de 2020].

World Health Organization. Coronavirus disease 2019 (COVID19) Situation Report –51. 11 março 2020. https://www.who.int/docs/default-source/coronaviruse/situation-reports/20200311-sitrep-51-covid-19.pdf?sfvrsn=1ba62e57_10 [acessado em 07 de abril de 2020].

World Health Organization. Coronavirus disease 2019 (COVID19) Situation Report –78. 07 April 2020.

https://www.who.int/docs/default-source/coronaviruse/situation-reports/20200407-sitrep-78-covid-19.pdf?sfvrsn=bc43e1b_2

